# Failure of Concentric Regulatory Zones to Halt the Spread of COVID-19 in South Brooklyn, New York: October-November 2020

**DOI:** 10.1101/2021.11.18.21266493

**Authors:** Jeffrey E. Harris

## Abstract

We relied on reports of confirmed case incidence and test positivity, along with data on the movements of devices with location-tracking software, to evaluate a novel scheme of three concentric regulatory zones introduced by then New York Governor Cuomo to address an outbreak of COVID-19 in South Brooklyn in the fall of 2020. The regulatory scheme imposed differential controls on access to eating places, schools, houses of worship, large gatherings and other businesses within the three zones, but without restrictions on mobility. Within the central red zone, COVID-19 incidence temporarily declined from 131.2 per 100,000 population during the week ending October 3 to 62.5 per 100,000 by the week ending October 31, but then rebounded to 153.6 per 100,000 by the week ending November 28. Within the intermediate orange and peripheral yellow zones combined, incidence steadily rose from 28.8 per 100,000 during the week ending October 3 to 109.9 per 100,000 by the week ending November 28. Data on device visits to pairs of eating establishments straddling the red-orange boundary confirmed compliance with access controls. More general analysis of device movements showed stable patterns of mobility between and beyond zones unaffected by the Governor’s orders. A geospatial regression model of COVID-19 incidence in relation to device movements across zip code tabulation areas identified a cluster of five high-mobility ZCTAs with estimated reproduction number 1.91 (95% confidence interval, 1.27-2.55). In the highly populous area of South Brooklyn, controls on access alone, without restrictions on mobility, were inadequate to halt an advancing COVID-19 outbreak.

## Introduction

The idea of drawing a series of concentric containment circles around an outbreak is well established in the control of communicable diseases. The U.S. Department of Agriculture, for example, has adopted the model of three concentric containment zones – the infected zone, the buffer zone, and the surveillance zone – as its standard practice to contain highly contagious animal diseases [1]. During the COVID-19 pandemic, the National Task Force in the Philippines established a four-circle scheme to enforce graded degrees of quarantine: a critical zone subject to complete lockdown, where a cluster of cases had been identified; a surrounding 500-meter-radius containment zone where a modified lockdown prevailed; a surrounding buffer zone subject to community-level quarantine; and surrounding outside area with further relaxation of mobility controls [2, 3].

In their classic form, concentric regulatory zones have served as quarantine boundaries, as one tool in the larger toolbox of strategies to restrict mobility in the control of infectious diseases [4]. Their use has been especially appealing when attack rates are directly related to the duration of contact and inversely related to the distance from an identifiable source, as they were in the Toronto-area SARS outbreak of 2003 [5]. It is well understood, however, that zone boundaries cannot simply be drawn around the areas of highest infection density, but need to take movement patterns into account [6].

On October 6, 2020, then New York Governor Cuomo issued a series of executive orders establishing a novel variation on the classic concentric control scheme [7]. Rather than serving as mass quarantine boundaries, the concentric areas would define the extent of access control to restaurants, schools, gyms, houses of worship, and large gatherings generally. While several areas of concern were identified throughout the state of New York, far and away the principal challenge was the surge of new COVID-19 cases in the South Brooklyn area of New York City.

Our task here is to combine data on COVID-19 incidence and testing outcomes with data on the movements of devices equipped with location-tracking software to evaluate what happened over the ensuing months. Our research approach follows a long line of studies of the role of mobility in the spread of contagion [8-10]. While the evidence shows that the Governor’s novel regulatory scheme failed to halt the surge of COVID-19, our focus here is on trying to understand exactly why.

### Background

By mid-September 2020, it had becoming increasingly evident that the recent surge of COVID-19 cases in certain hotspots of New York City was threatening the city’s reopening plans. By September 29, then New York City Mayor de Blasio had signaled his intention to close nonessential businesses and all public and private schools in nine zip codes in the boroughs of Queens and Brooklyn for 14-28 days [11]. The target zip codes included five in Brooklyn: Borough Park (11219), Gravesend (11223), Midwood (11230), Bensonhurst (11204), Flatlands (11210), and Gerritsen Beach/Homecrest/Sheepshead Bay (11229). Test positivity rates had increased beyond the acceptable threshold of 3% in these areas, exceeding 7% in Gravesend (11223) [12].

On October 6, however, then New York State Governor Cuomo intervened with his own regulatory control strategy, which he termed a “cluster action initiative.” [7] Developed in consultation with public health experts, the initiative imposed new local restrictions on activity within “red zones” where clusters of new cases had been identified [13, 14]. Recognizing that individuals within these high-risk areas tended to “interface with the surrounding communities,” the initiative established two concentric rings – an intermediate orange zone and a peripheral yellow zone – surrounding the high-risk red zone.

Zone boundaries were drawn based on the test positivity rate, that is, confirmed COVID-19 cases as a percentage of all persons presenting for testing. Among highly populated areas, which included the borough of Brooklyn, a red zone was defined as having a sustained test positivity rate above 4 percent, while an orange zone had a rate from 3 to 4 percent, and a yellow zone had a rate from 2.5 to 3 percent [13].

Among the restrictions on activity, a red zone prohibited mass gatherings, allowed only essential businesses to open, closed in-person schooling, and restricted restaurants and other food providers to takeout/delivery only. An orange zone allowed gatherings up to 10 people, closed only high-risk businesses such as gyms and personal care, closed in-person schooling, and allowed outdoor dining with up to 4 persons per table. A yellow allowed gatherings up to 25 people, permitted all businesses to open, permitted indoor as well as outdoor dining up to 4 persons per table, and opened schools to in-person instruction subject to mandatory testing of students, teachers and staff [13]. Restrictions on access to houses of worship were also initially imposed, with limits of 25% capacity in a red zone, 33% capacity in an orange zone, and 50% capacity in a yellow zone, but were subsequently blocked by the U.S. Supreme Court [15]. The sanctions for failure to comply included withholding of funds to localities and schools [16].

Revisions of zone boundaries and changes in zone classification were based principally on the test positivity rate. On October 21, the Governor, citing the early success of the strategy in Brooklyn, reclassified the borough’s original orange zone as a yellow zone, while the red zone remained unchanged [14]. On November 3, citing further gains, the Governor reduced the size of the red zone by half [17]. A few days later, on November 9, the red zone was reclassified as an orange zone [17], and subsequently as a yellow zone on November 18 [18]. The zones were eventually dissolved without fanfare in January 2021.

## Materials and Methods

### Data Sources: Regulatory Zone Boundaries

We determined regulatory zone boundaries from detailed maps issued by the office of the Governor of New York [19-21], along with accompanying announcements of updates [17, 18]. Fig. A1 in Supplement Appendix A shows the boundaries of the original concentric red, orange, and yellow zones, effective October 9, 2020, overlaid on a street map of the larger New York City area [19]. Figs. 1a and 1b below depict the evolution of the regulatory zone boundaries, overlaid on more detailed street maps of South Brooklyn.

**Fig 1.**
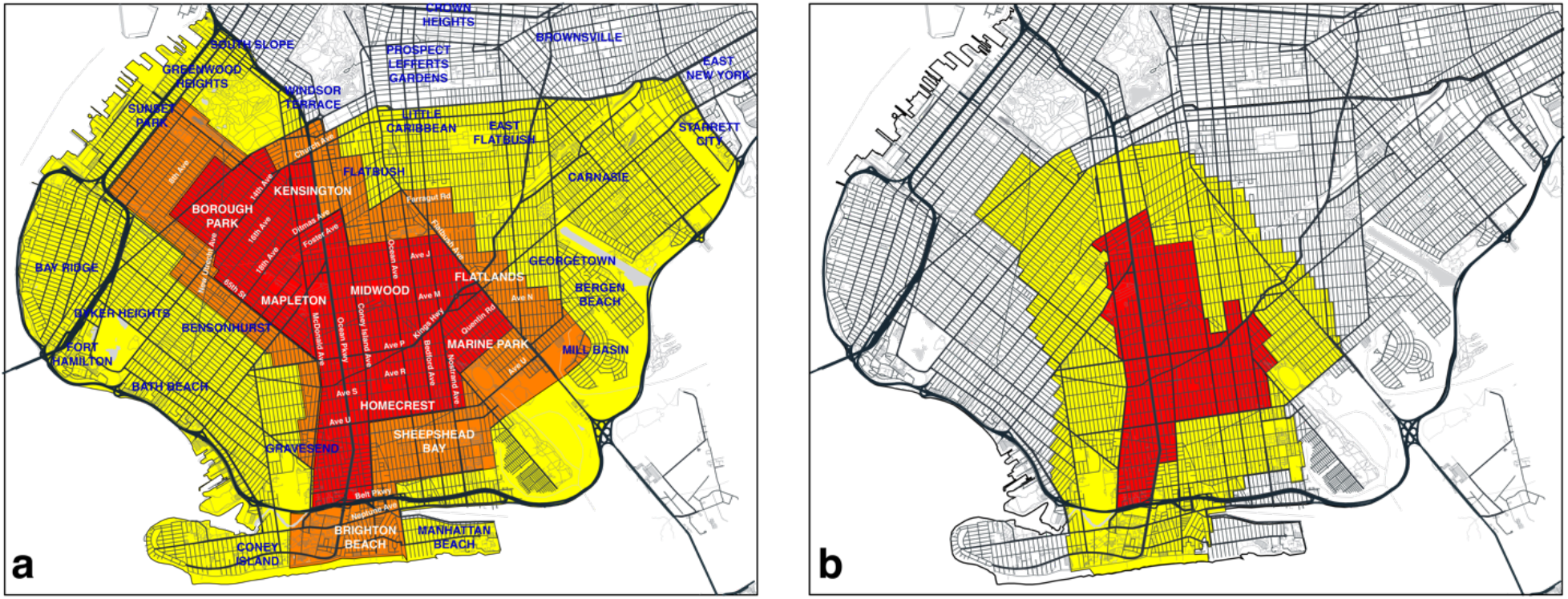
**a. Red, Orange and Yellow Zones Effective October 9, 2020 [19], Overlaid on Annotated Map of South Brooklyn**. On October 21, the orange zone was redesignated as a yellow zone, while the red zone remained unchanged [20]. **b. Revised Boundaries of Red and Yellow Zones as of November 3 [22]**. On November 9, the red zone was redesignated as an orange zone [17], and subsequently as a yellow zone on November 18 [18]. For base map of New York City streets, see [23].

Fig. 1a identifies the original red, orange and yellow zones [19]. On October 21, the orange zone was incorporated into the existing yellow zone, while the original red zone boundaries remained unchanged [20]. Fig. 1b shows the contracted red and yellow zones as of November 3 [21]. On November 9, the red zone was redesignated as an orange zone [17], and subsequently as a yellow zone on November 18 [18].

### Data Sources: Zip Code Tabulation Area Boundaries

Figs. 2a and 2b superimpose the respective regulatory boundaries of Figs. 1a and 1b on a map of zip code tabulation areas (ZCTAs) in South Brooklyn. The lack of complete congruence between the ZCTA and regulatory boundaries is evident. As discussed below, geographically detailed data on confirmed COVID-19 incidence over time was available only at the ZCTA level. Accordingly, for the purposes of analyzing COVID-19 incidence, we classified any of the nine ZCTAs that even partially overlapped the original red zone as an original red-zone ZCTA. These ZCTAs, indicated in boldface in Fig. 2a, included 11204, 11210, 11218, 11219, 11223, 11229, 11230, 11234, and 11235. The remaining 12 ZCTAs, indicated in italics, were classified as original orange-yellow zone ZCTAs. In Fig. 2b, the same classification of ZCTAs is shown in the map of the contracted regulatory zones effective November 3.

**Fig 2.**
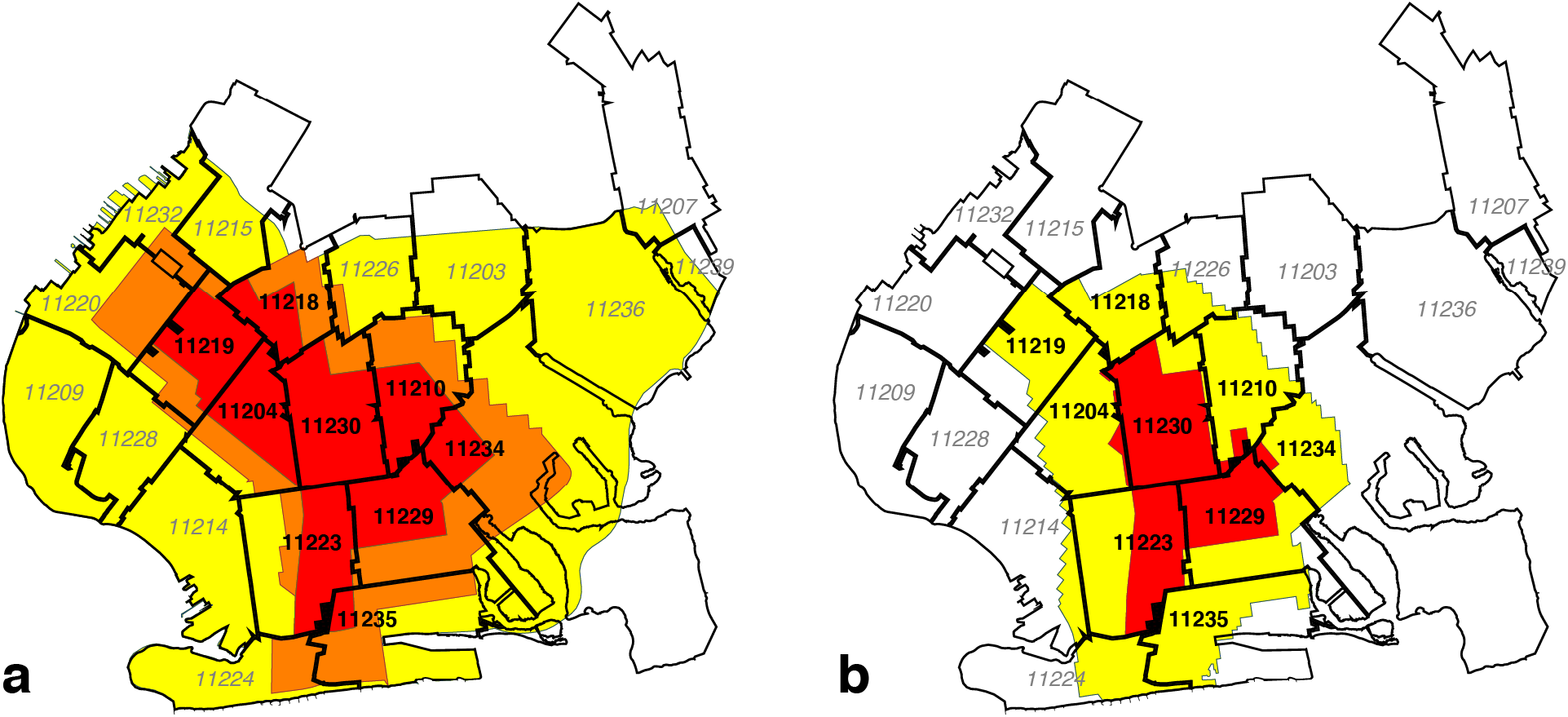
**a. Boundaries of the Original October 9 Zones Overlaid on Map of Zip Code Tabulation Areas (ZCTAs)**. In our calculations of confirmed COVID-19 incidence rates and test positivity rates, we assigned the nine ZCTAs in black boldface to the original red zone, while the remaining 12 ZCTAs in gray italic were assigned to the combined orange and yellow zones. **b. Updated Boundaries of the Redrawn November 3 Zones Overlaid on the Same ZCTA Map**. For base map of New York City ZCTAs, see [24].

### Data Sources: Census Block Groups

As discussed below, we relied on the Safegraph Social Distancing database [25] to gauge the movements of devices equipped with location-tracking software throughout the greater New York City area. The origin and destination of each device movement in the Social Distancing database are keyed to census block groups (CBGs). Accordingly, we developed a separate correspondence between CBGs and regulatory zones, as shown in Fig. 3a. Relying on QGIS software [26], we determined the geocoordinates the centroids of all CBGs in South Brooklyn based upon their U.S. Census-defined shape files [27]. Using the Stata routine *geoinpoly* [28], we then assigned each CBG to the regulatory zone polygon in which its centroid was situated. While not explicitly shown in the figure, we used the same procedure to map CBGs into the ZCTA polygons shown in Figs. 2a and b.

**Fig 3.**
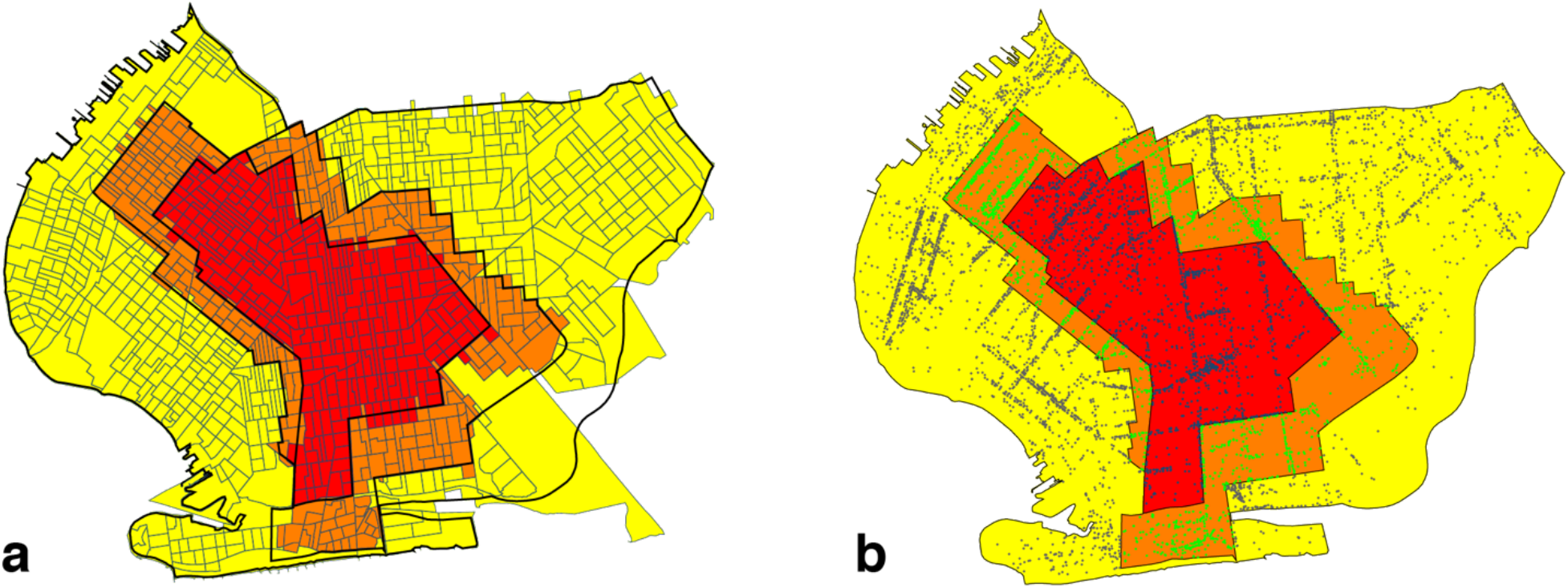
**a. Boundaries of the Original October 9 Zones Overlaid on Map of Census Block Groups (CBGs).** In our calculations of within- and between-zone device movements, we assigned each CBG to the zone containing its geographic centroid. **b. Boundaries of the Original October 9 Zones Overlaid on Map of Safegraph Points of Interest (POIs)** [29]. Each point corresponds to a POI. Points have different colors depending on the regulatory zone in which they were located. For base map of New York City census block groups, see [30].

### Data Sources: Points of Interest

We relied upon the SafeGraph Patterns database [29] – a source distinct from the Social Distancing database [25] – to analyze visits of devices to points of interest (POIs) within the regulated area in South Brooklyn. At the broadest level, Safegraph classifies POIs according to the variable *top_category*, which includes such categories as “Automotive Repair and Maintenance,” “Child Day Care Services,” “Clothing Stores,” “Elementary and Seconday Schools,” “Gasoline Stations,” and “Health and Personal Care Stores.” One of the largest such categories is “Restaurants and Other Eating Places.” Taking advantage of the Safegraph-supplied geocoordinates of each POI, and again relying on the Stata *geoinpoly* routine [28], we classified each POI as being located in one of the original three regulatory zones. Based upon this classification, we constructed Fig. 3b, which plots the location of every POI as a color-coded point within the original October 9 regulated area.

### Data Sources: COVID-19 Incidence and Test Positivity

We relied upon data published by the New York City Department of Health on COVID-19 incidence, measured as the number of confirmed cases per 100,000 population, and COVID-19 test positivity, measured as the percentage of positive tests, broken down by ZCTA and week [31, 32]. Both data sources covered the weeks ending August 8 through November 28, 2020.

### Statistical Methods: Paired Point-of-Interest Analysis

We developed a paired point-of-interest (POI) analysis to test whether the regulations imposed by the Governor were in fact enforced and effective. To that end, we focused on device movements into restaurants and other eating places located in the original red zone, where establishments were restricted to takeout/delivery only, and in the original orange zone, where establishments could also offer outdoor dining with up to 4 persons per table [13]. To avoid potentially biased comparisons between local dining patterns in distinct neighborhoods (such as Borough Park in the original red zone and Brighton Beach in the original orange zone, as shown in Fig. 1a), we restricted our comparisons to pairs of nearby eating establishments that straddled the original red-orange border.

We identified all POIs in the SafeGraph Patterns database [29] with a *top_category* designated as “Restaurants and Other Eating Places.” Within this restricted dataset, there were 395 POIs in the original red zone and 507 POIs in the original orange zone. (There were also 1,045 POIs in the original yellow zone, but they were not included in our paired POI analysis.) Relying on the Stata program *geonear* [33], we isolated 219 pairs of red-zone and orange-zone POIs that were nearest neighbors of each other, where the maximum distance between POIs within each pair was 300 meters. The median distance between paired POIs was 130.9 meters, with 25^th^ and 75^th^ percentiles equal to 70.7 and 218.6 meters, respectively. Because a POI on one side of the red-orange boundary could be the nearest neighbor of multiple POIs on the other side, the resulting dataset contained 145 unique red-zone POIs and 114 unique orange-zone POIs. Supplement Appendix Fig. A2 maps two such pairs straddling the red-orange boundary running along Avenue U in Brooklyn.

Let *I* denote the set of all red-zone POIs, with typical element *i* ∈ *I*, and let *J* denote the set of all orange-zone POIs, with typical element *j* ∈ *J*. Then our database consists of a subset of 219 unique pairs *(i, j)* contained within the larger set *I* × *J*. For each POI, we relied on the Safegraph Patterns variable *visits_by_day* to compute the number of visits during each week, starting with the week ending October 1 (designated *t* = 0) and continuing through the week ending December 3 (*t* = 9). We thus had 219 paired observations (*y*_*Rit*_, *y*_*Ojt*_), where *y*_*Rit*_ represents the number of visits during week *t* to red-zone POI *i*, and where *y*_*Ojt*_ represents the number of visits during week *t* to orange-zone POI *j*.

Given these data, we ran the following fixed-effects regression model:

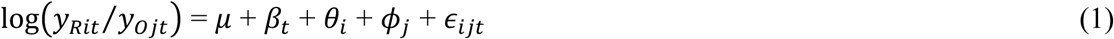

In equation (1), the parameter *μ* is an overall constant term, *β*_*t*_, *θ*_*i*_. and *ϕ*_*j*_ are fixed-effect parameters corresponding to each week *t*, red-zone POI *i*, and orange-zone POI *j*, and *∈*_*ijt*_ are independently distributed spherical error terms. Within this fixed-effects framework, only the contrasts *β*_*t*_ – *β*_0_ (*t* = 1,…,9) can be identified. If the regulations imposed by the Governor were in fact enforced and effective, then we would expect the estimated parameters *β*_*t*_ to be negative.

### Statistical Methods: Geospatial Analysis

Movements to specific points of interest such as restaurants, auto repair shops and daycare centers are part of a larger set of movements to destinations that include private residences and workplaces. We sought to determine how these more general mobility patterns related to the evolution of COVID-19 incidence, particularly during the period from the second half of October to the end of November, when cases of the disease were increasing throughout the regulated area in South Brooklyn. To that end, we developed a geospatial model relating COVID-19 incidence to general device movements. The central feature of this model was that the incidence of the disease in a particular ZCTA during a particular week was related to the incidence in all ZCTAs during the prior week. Moreover, the influence of one ZCTA on another was determined by the volume of device traffic between the two. The details of our model and its implementation are given in Supplement Appendix B.

Our model relied on two types of data: COVID-19 incidence and device movements. Because our data on COVID-19 incidence were based upon ZCTAs, we classified device movements between ZCTAs as well. Relying on data published by the New York City Department of Health [31], we constructed a data series {*y*_*kt*_} of the incidence of confirmed COVID-19 cases per 100,000 population in ZCTA *k* during week *t*, where *k* = 1,…,21 indexes the 21 ZCTAs within the regulated area in Fig. 2, and where *t* = 1,…,7 indexes the 7-week period running from the week ending October 17 through the week ending November 28. Relying upon the variables *origin_census_block_group* and *destination_cbgs* in the Safegraph Social Distancing database [25], we constructed a data series {*n*_*kℓt*_} of counts of device movements from ZCTA *k* into ZCTA *ℓ* during week *t*. The counts *n*_*kkt*_, which represented the number of device movements staying within ZCTA *k* during week *t*, included those devices homed the ZCTA that made no movements. While we also observed device movements beyond the 21-ZCTA regulated area, as well as movements into the regulated area from outside, we focused sharply on the regulated area in order to ascertain how the traffic between local ZCTAs influenced the dynamics of COVID-19 transmission.

Based upon our underlying data on device movements, we let *V*_*t*_ be a 21×21 square matrix with typical element *ν*_*kℓt*_ measuring the proportion of all devices originating in ZCTA *k* that moved into ZCTA *ℓ* during week *t*. The elements of each row of *V*_*t*_ sum to 1. Likewise referring to week *t*, we let *W*_*t*_ be a 21×21 square matrix with typical element *w*_*kℓt*_ measuring the fraction of all devices with a destination in ZCTA *ℓ* that originated in ZCTA *k*. The elements of each column of *W*_*t*_ sum to 1. Let *Y*_*t*_ denote a 21×1 column vector with elements *y*_*kt*_. We let *Y*_*t*+1_ represent the corresponding vector of incidence rates one week later, while *η*_*t+1*_ is a contemporaneous vector of error terms. Under the strong assumption of *homogeneous mixing*, our geospatial model yields:

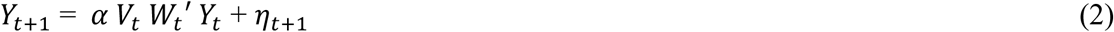

The model of equation (2) allows for a susceptible individual homed in ZCTA *k* to be infected not only through contact with another resident of the same ZCTA *k*, but also through contact with a resident of another ZCTA *ℓ*. The inclusion of both vectors *V*_*t*_ and *W*_*t*_, moreover, allows for two loci of transmission from an infected individual residing in ZCTA *ℓ* to a susceptible individual homed in ZCTA *k*. Either the susceptible individual homed in *k* had temporarily moved to *ℓ*, or the infected individual in *ℓ* had temporarily moved to *k*. The unknown parameter *α* in equation (2), to be estimated from the data, represents the uniform reproductive number for COVID-19 transmission throughout the entire regulated area for the 7-week time period under study.

### Inhomogeneous Mixing

The assumption of homogeneous mixing, with a uniform reproductive number *α*, is strong. Accordingly, we considered two alternative specifications involving inhomogeneous mixing. First, we assumed instead that movements by individuals who remained within their home ZCTA could have a different influence on COVID incidence. To capture the effect of these within-ZCTA device movements, we defined *D*_*t*_ = diag(*V*_*t*_ *W*_*t*_′) as the *K* × *K* square matrix with the same diagonal elements as *V*_*t*_*W*_*t*_′ but zero off-diagonal elements, and then introduced the additional regressor *D*_*t*_*Y*_*t*_ into our model. Defining the *K* × *K* square matrix *X*_*t*_ = *V*_*t*_*W*_*t*_′ − *D*_*t*_, we have:

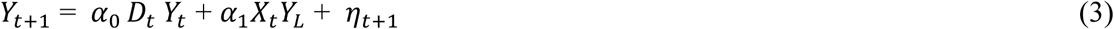

We refer to this alternative as *inhomogeneous mixing model A*. In equation (3), the parameter *α*_0_ reflects the reproductive number for within-ZCTA movements, while the parameter *α*_1_ reflects the corresponding reproductive number for between-ZCTA movements. We estimated the models of equation (2) with weighted least squares, where the weights were the ZCTA populations derived from the New York City Department of Health data [31].

Second, we relaxed the assumption that all between-ZCTA movements had the same reproductive number *α*_1_. Instead, movements to and from certain high-risk ZCTAs were permitted to exert more influence than movements to and from the remaining lower-risk ZCTAs. To capture such differences in transmission efficiency, we partitioned the set of ZCTAs into two mutually exclusive subsets, *L* and *H*, representing the low- and high-transmission ZCTAs respectively. Conformally partitioning *X*_*t*_ vertically into two matrices, *X*_*tL*_ and *X*_*tH*_, and the column vector *Y*_*t*_ horizontally into two vectors: *Y*_*tL*_ and *Y*_*tH*_, our equation (3) becomes:

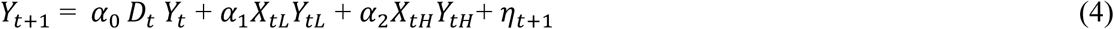

The unknown parameters *α*_0_, *α*_1_, and *α*_2_, respectively, represent the reproductive numbers for movements within-ZCTAs, movements to and from low-risk ZCTAs, and movements to and from high-risk ZCTAs. We refer to this alternative as *inhomogeneous mixing model B*.

We performed two tests of inhomogeneous mixing model B, based upon two different partitionings of the set of 21 ZCTAs delineated in Fig. 2. First, on the basis of our examination of the trends in inter-ZCTA mobility, as detailed in the Results section, we identified five high-mobility ZCTAs along the southern boundary of the 21-ZCTA area as the most likely elements of the high-risk set *H*. The estimates based upon this high-low partitioning of ZCTAs were identified as B1. Second, we relied on the original classification of regulatory zones specified in the Governor’s order of October 6, with the red-zone ZCTAs specified as high risk (*H*) and the remaining orange and yellow zones specified as low risk (*L*). These estimates were identified as B2. We similarly estimated the inhomogeneous geospatial models of equations (3) and (4) with population-weighted least squares.

## Results

### COVID-19 Incidence and Test Positivity

Fig. 4 below plots the incidence of confirmed cases of COVID-19 per 100,000 population in the original red zone, the original combined orange and yellow zones, and in the rest of New York City during the weeks ending August 8 through November 28. Also noted in the plot are the dates of Governor’s five successive regulatory actions, starting with the imposition of the original three concentric zones, effective October 9. While the regulatory zones underwent revisions, the geographic areas used to compute case incidence in Fig. 4 remained unchanged.

**Fig. 4.**
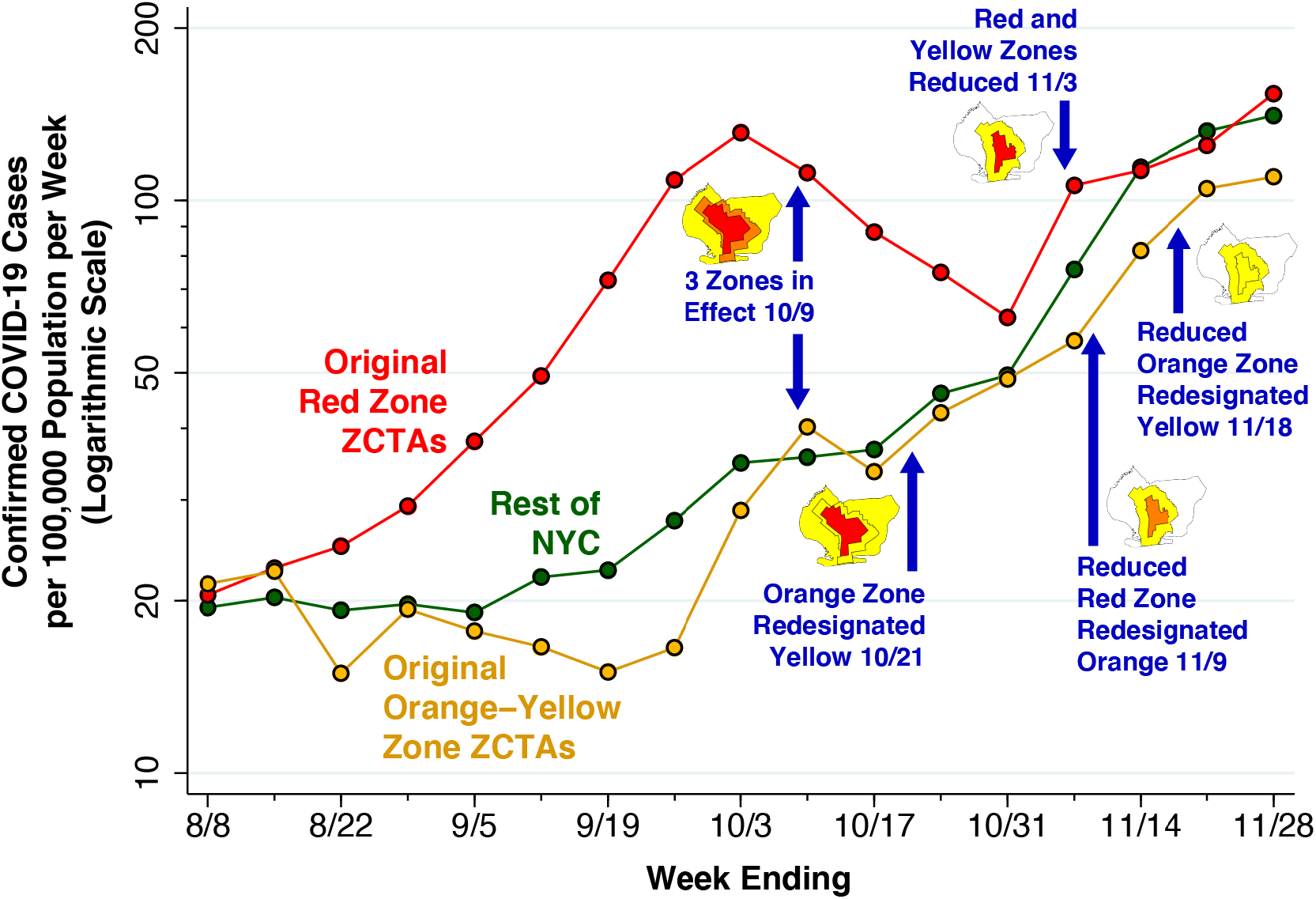
Confirmed COVID-19 Cases per 100,000 Population in the Original Red Zone, Original Combined Orange-Yellow Zones, and the Rest of New York City, Weeks Ending 8/8 Through 11/28/2020. The data points represent population-weighted rates, derived from ZCTA-specific weekly COVID-19 incidence as reported by the NYC Department of Health [31], and aggregated according to the scheme in Fig. 2a. Superimposed on the graphic are the dates of five successive regulatory actions: (i) establishment of the original three zones on October 9 [7, 19]; (ii) redesignation of the original orange zone as yellow on October 21 [14]; (iii) reduction in the size of the red and orange zones on November 3 [17, 22]; (iv) redesignation of the reduced red zone as orange on November 9 [17]; and (v) redesignation of the reduced orange zone as yellow on November 18 [18].

After rising during August and September, COVID-19 incidence in the ZCTAs comprising the original red zone started to decline during the week ending October 10, a time period that included five days before the regulatory scheme took effective. COVID-19 incidence in the red zone continued to decline through the week ending October 31, but then began to rebound. By contrast, COVID-19 incidence in the original orange and yellow zones, as well as the rest of New York City, had been increasing since at least mid-September, and reached approximately the same level as the original red zone by November. By November 21, all three series had exceeded the threshold of 100 cases per 100,000 population per week.

Fig. 5 focuses sharply on the original red zone. The incidence of confirmed COVID-19 cases per 100,000 population, measured on the left-hand vertical scale, is reproduced from Fig. 4. Superimposed on this time series is the test positivity rate, measured on the right-hand vertical scale. As in Fig. 4, the dates when each of the five successive regulatory actions went into effect are noted. Again, while the regulatory zones underwent successive revisions, the geographic area used to compute the positivity rate – namely the original red zone – remained unchanged.

**Fig. 5.**
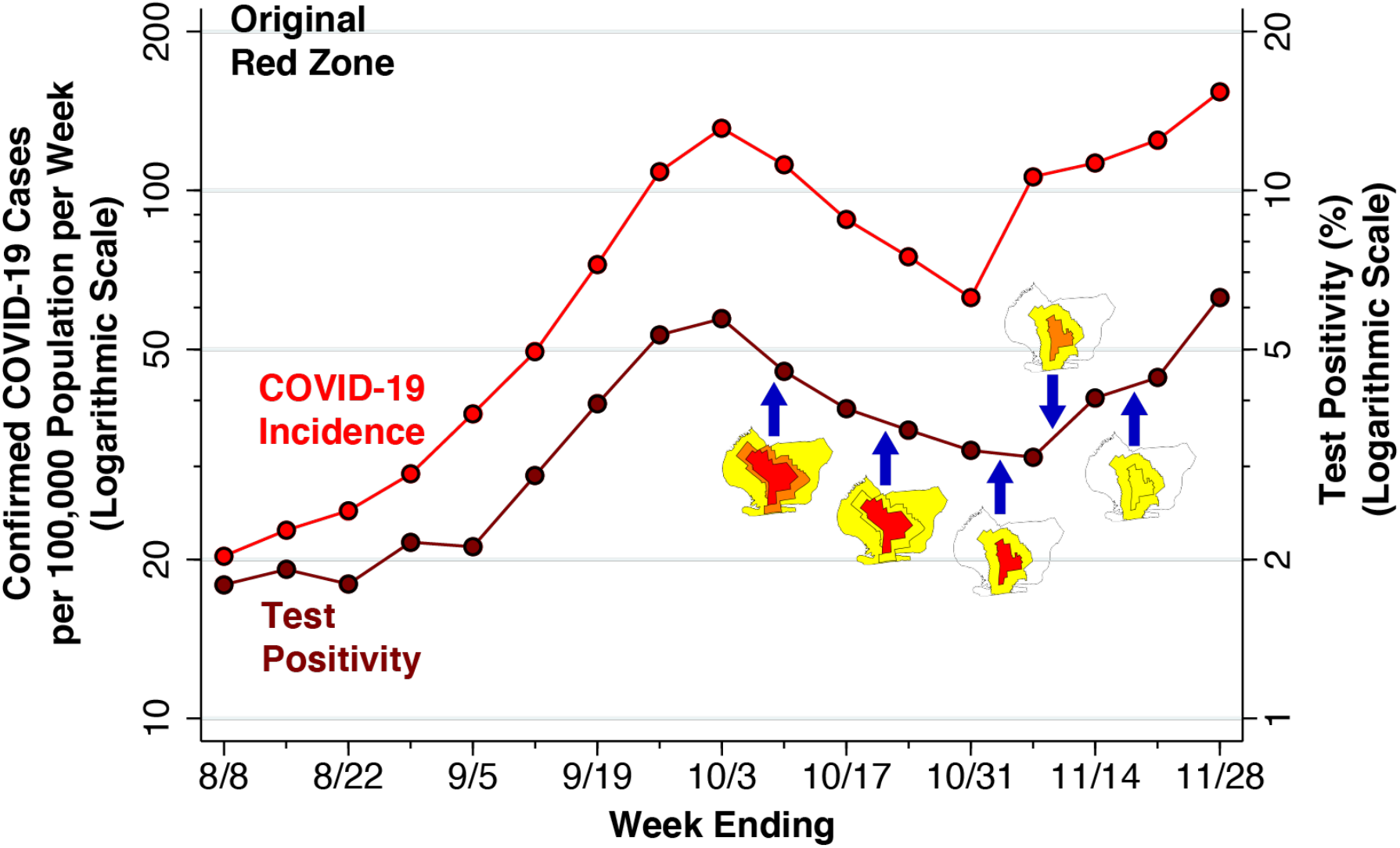
Confirmed COVID-19 Cases per 100,000 Population (Left Axis) and Test Positivity Rate (Right Axis) in the Original Red Zone, Weeks Ending 8/8 Through 11/28/2020. As in Fig. 4, the data points represent population-weighted rates, derived from ZCTA-specific weekly COVID-19 incidence [31] and test positivity [32] among the ZCTAs identified as covering the red zone in Fig. 2a. As in Fig. 4, the blue arrows show the dates on which the five successive regulatory actions went into effect, while the accompanying maps show the corresponding zone changes.

The variable vertical gap between the two timeseries in Fig. 5 corresponds to the changing testing rate for COVID-19. Thus, the testing rate per 1,000 population progressively increased from 11.33 during the week ending August 8 to 17.15 during the week ending September 12, and then further increased to 24.61 by the week ending October 10. By the weeks ending November 7, 14 and 21, respectively, the testing rates had reached 33.99, 27.88, and 28.20 per 1,000 population.

Because the red-zone ZCTAs in Fig. 2a only approximate the precise boundaries of the original red zone, the estimated test positivity rates plotted in Fig. 5 represent only approximations to the positivity rates that were relied upon by state regulators. Still, during the week ending November 7, there is a striking divergence between the declining test positivity rate and the concurrently rising incidence rate. This finding suggests that regulators, relying on the trend in a test positivity rate that was biased downward by enhanced testing, relaxed restrictions when in fact the incidence of the disease was rising.

Fig. 6 below tracks the detailed evolution of COVID-19 incidence in each of the 21 ZCTAs identified in Fig. 2 during the weeks ending September 12 through November 28. The first row, covering the weeks ending September 12 through October 3, shows increasing disease incidence in the central ZCTAs covering what would ultimately be designated as the red zone. In the second row, covering the weeks ending October 10 – 31, the incidence of COVID-19 was declining in the central ZCTAs but increasing in the peripheral ZCTAs, particularly along the southern and western borders of the South Brooklyn region. In the third row, covering the weeks ending November 7 – 28, COVID-19 incidence continued to rise in these peripheral ZCTAs, while resuming its upward trend in the central ZCTAs. By November 28, the original central zone of high-incidence ZCTAs is no longer distinguishable.

**Fig. 6.**
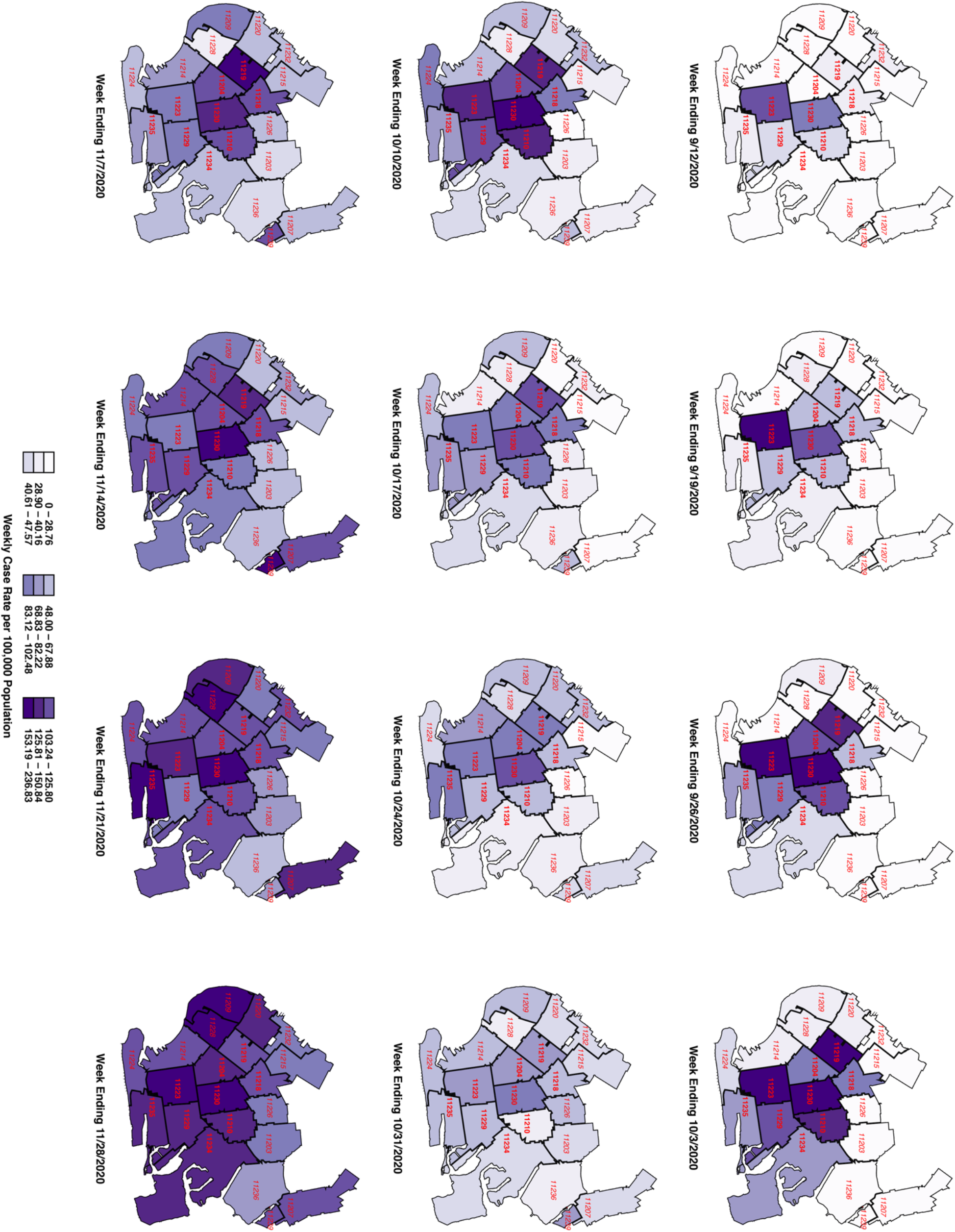
Evolution of Weekly Case Rate per 100,000 Population Among 21 ZCTAs in South Brooklyn, Weeks Ending 9/12 Through 11/28/2020. For base map of New York City ZCTAs, see [24].

### Visits to Restaurants: Paired POI Analysis

Fig. 7 below shows the results of our paired POI analysis of visits to restaurants and other eating places. The estimate of *β*_1_ = –0.075 for the week ending October 8 is negative and significantly different from zero in a two-sided test (p = 0.0497). That is, visits to restaurants in the red zone had already declined by 7.5% relative to those in the orange zone during the week before the regulatory scheme went into effect. While the individual estimates of *β*_2_ = –0.059 and *β*_3_ = –0.067 are not significantly different from zero, the overall downward trend is evident by the weeks ending October 29 and November 5, where *β*_4_ = –0.114 (p = 0.003) and *β*_5_ = –0.172 (p < 0.001). Thereafter, as the red zone is reduced by half (effective November 3), then reclassified as orange (effective November 9), and then reclassified as yellow (November 18), the estimates of *β*_*t*_ begin to rebound. By the week ending December 3, the estimate *β*_9_ = –0.006 is no longer significantly different from zero (p = 0.87).

**Fig. 7.**
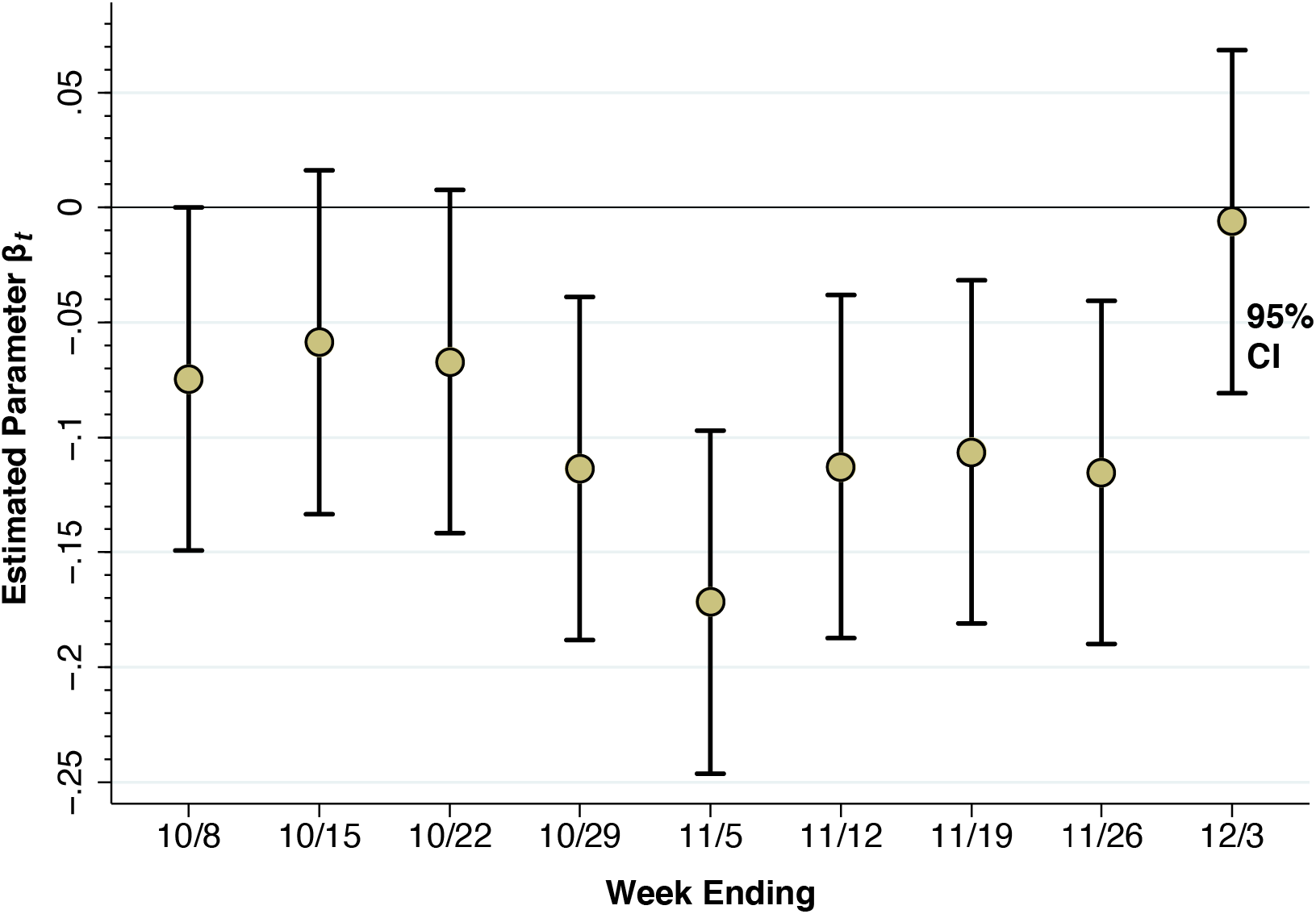
Estimated Time-Specific Fixed Effects Derived from the Paired Point-of-Interest Analysis. Each point represents an estimate of the contrast *β*_*t*_ – *β*_0_ for *t*=1,…,9, where *t* = 0 represents the week ending 10/1/20 as a reference category. The error bars surrounding each point represent 95% confidence intervals.

### Movements Within and Between Regulatory Zones

The results of our paired analysis of restaurants and other eating places, as shown in Fig. 7, narrowly reflect trips to a specific category of establishments that were subject to specific regulatory controls. They do not necessarily capture broader trends in unregulated movements of individuals within and between zones.

Supplement Appendix Table A1 delineates movements of devices within and between zones, as well as movements outside the regulated area, during the three weeks before and the subsequent three weeks after the regulations went into effect on October 9. Comparison of the movement matrices during the two time periods (September 18 through October 8, October 9 through October 29) indicates that overall movement patterns remained stable. Among devices homed in the original red zone, only 57–58% of movements were confined to the red zone, while 23–24% of movements were to destinations outside the regulated area entirely. The same pattern is evident in the movements of devices homed in the original orange and yellow zones as well.

The overall stability of within-zone movements seen in Supplement Appendix Table A1 could still obscure significant changes in mobility, particularly in the proportion of devices that made few if any movements. Supplement Appendix Fig. A2, however, shows that the percentage of devices that made no movements at all rose only by 1 to 2 percentage points.

Fig. 8 tracks more detailed movements between ZCTAs, rather than between regulatory zones, during October 25 through November 28. This time period corresponds to the final five weekly maps in Fig. 6, when COVID-19 was increasing both in the central ZCTAs and the peripheral ZCTAs along the southern and western edges of the regulated area. The width of each arrow corresponds to the magnitude of the flow between ZCTAs. The figure demonstrates that dominant inter-ZCTA movements were between four red-zone ZCTAs (11229, 11223, 11230, 11204) and three orange- and yellow-zone ZCTAs (11235, 11224, 11214). Moreover, there was significant device traffic between these southern ZCTAs and other peripheral ZCTAs to the west of the regulated area.

**Fig. 8.**
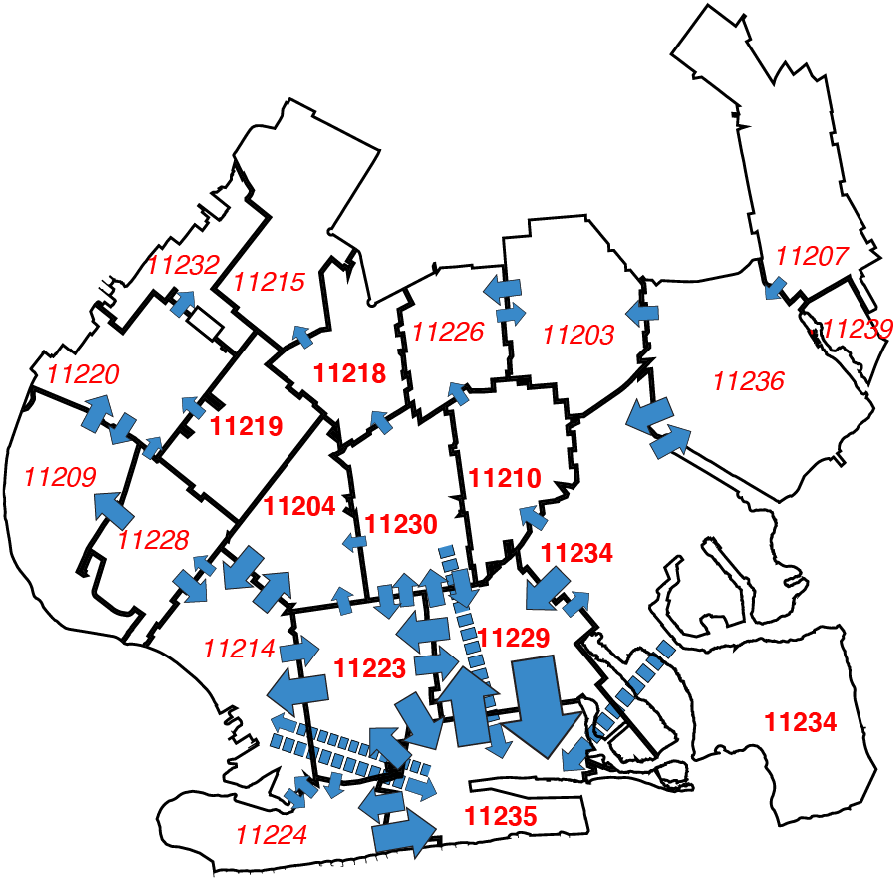
Most Frequent Transits Between ZCTAs During 10/25 Through 11/28/2020. The width of each blue arrow is proportional to the number of device movements in the Safegraph cohort. The longer arrows with dashed lines represent transits between noncontiguous ZCTAs. The number of device movements ranged from 21,139 (corresponding to the transit 11229 → 11235) to 5,002 (corresponding to 11214 → 11224). Transits between ZCTAs with fewer than 5,000 movements are not shown. The transits captured by the arrows comprised 44.5% of all device movements between ZCTAs. For base map of New York City ZCTAs, see [24].

### Influence of High-Risk ZCTAs: Geospatial Analysis

Based upon our findings in Fig. 8, we identified five high-mobility ZCTAs as potential candidates for high-risk transmission to other surrounding ZCTAs in the area: 11223, 11229, 11235, 11224. and 11214. As seen in Fig. 2a, two of the areas, 11224 and 11214, were not in the original red zone, while another area, 11235, was excluded from the red zone when the zones were contracted on November 3.

Fig. 9 shows the principal results of our geospatial analysis. At the left end of the graphic, in a model of homogeneous mixing with a single, uniform parameter for the entire regulated area (equation 2 in the Methods section), the estimated reproductive number was *α* = 1.157 with 95% confidence interval [1.106, 1.208]. The middle panel shows the results of a two-parameter model of inhomogeneous mixing A, which allowed for movements within ZCTAs to have a different effect on COVID-19 transmission (equation 3). Both parameters *α*_0_ for within-ZCTA movements and *α*_1_ for between-ZCTA movements were in the range of 1.1–1.2, though they were too imprecise to be distinguishable from each other or from 1.0.

**Fig. 9.**
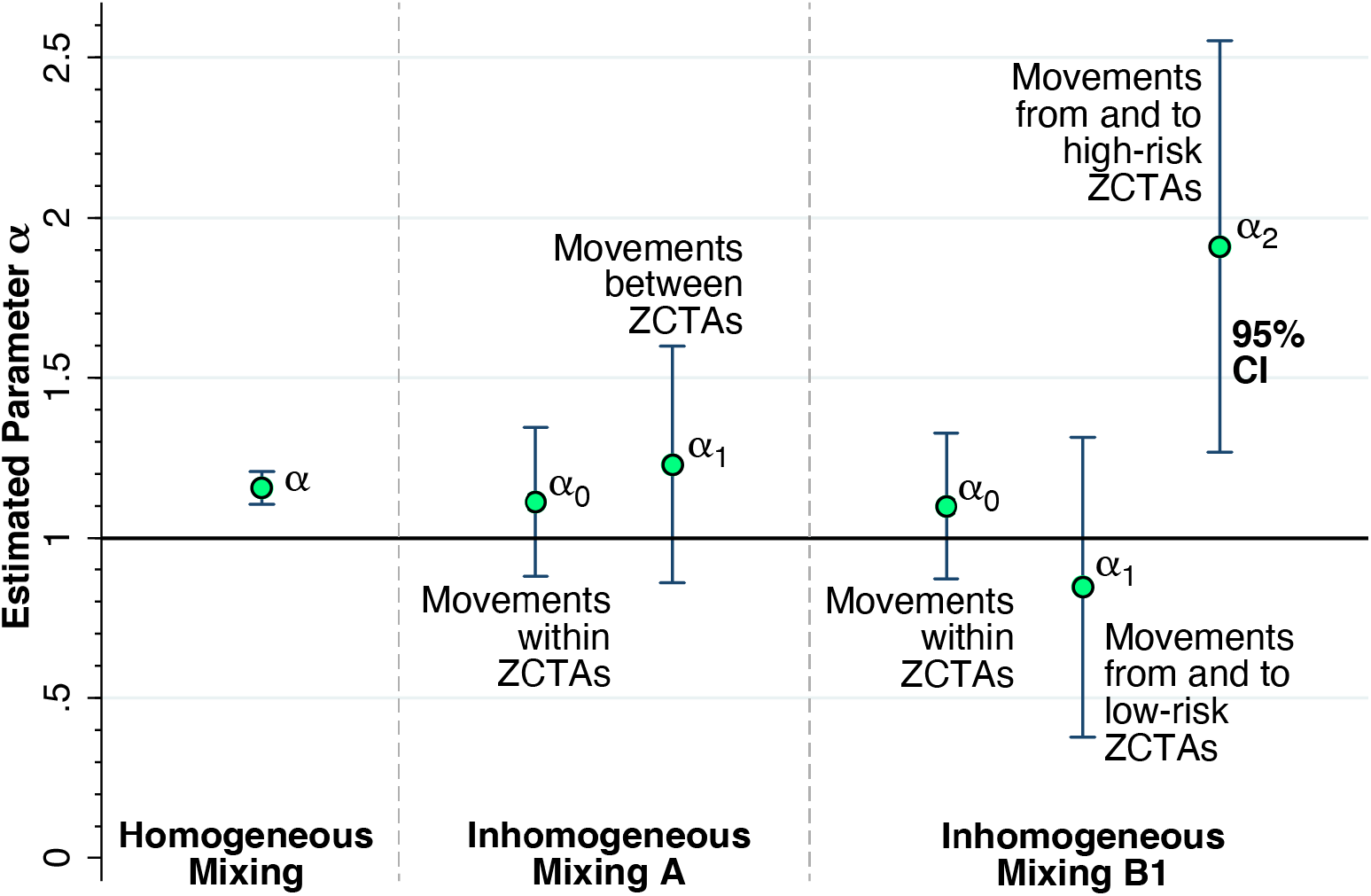
Estimates of the reproductive number *α* from a series of geospatial analyses. Left: Estimate of a uniform parameter *α* = 1.157 with 95% confidence interval [1.106, 1.208] for the entire regulated region, based upon the assumption of homogeneous mixing. The estimate of *α* is significantly different from 1.0 (2-sided F-test, p < 0.001). Middle: Estimates of the parameters *α*_0_ for the effect of movements within ZCTAs and *α*_1_ for between-ZCTA movements, based upon a two-parameter model of inhomogeneous mixing A. Right: Estimates of the parameters *α*_0_ for the effect of movements within ZCTAs, *α*_1_ for the effect of movements from and to low-risk ZCTAs, and *α*_2_ for the effect of movements from and to high-risk ZCTAs 11223, 11229, 11235, 11224, and 11214, based upon a three-parameter model of inhomogeneous mixing B1. The estimated parameter *α*_2_ = 1.910 with 95% confidence interval [1.268, 2.552] was significantly different from 1.0 (2-sided F-test, p = 0.012). Full details are given in Supplement Appendix Table C1.

The right-hand panel of Fig. 9 shows the estimates of the three-parameter model with inhomogeneous mixing B1 (equation 4), which further distinguished the five high-risk ZCTAs 11223, 11229, 11235, 11224, and 11214 from the remaining sixteen low-risk ZCTAs. The estimate of *α*_0_ was imprecise but consistent with the findings in the two-parameter model A in the middle panel. The estimate of *α*_1_ for low-risk ZCTAs was likewise imprecise, but pointed to a reproductive number less than 1.0. By contrast, the estimate of *α*_2_ for the effect of device traffic to and from the five high-risk ZCTAs gave a reproductive number of 1.910 with 95% confidence interval [1.268, 2.552]. Moreover, the estimate of *α*_2_ was significantly different from *α*_0_ (p = 0.045) and *α*_1_(p = 0.012) based upon 2-sided F-tests.

The alternative test of the three-parameter model with inhomogeneous mixing B2 (equation 4), based upon the classification of all the original red-zone ZCTAs as high risk, did not yield such precise results. The estimate of *α*_2_ for the effect of device traffic to and from these original red-zone ZCTAs gave a reproductive number of 1.485 with 95% confidence interval [0.992, 1.978]. This estimate was not significantly different from *α*_0_ (p = 0.284) of from *α*_1_ (p = 0.124). The complete results of the geospatial analysis are shown in Supplement Appendix Table C1.

## Discussion

### Non-identifiability of the Effect of the Governor’s Regulatory Scheme

Our analysis of paired eating places straddling the red-orange border suggests that the Governor’s October 6 regulatory scheme did indeed have some effect. The red-zone rules allowed for only takeout and delivery, while the less stringent orange-zone rules allowed for outdoor dining as well [13]. By the week ending November 5, as seen in Fig. 7, device visits to establishments on the red side of the red-orange border were down 17.2 percent more than their counterparts on the orange side. Attributing the entire decline to the regulatory scheme is problematic, however, inasmuch as device visits to establishments on the red side of border were already dropping more rapidly during the week ending October 8. Aside from voluntary action on the part of the red zone’s residents, the Mayor’s threats to impose controls in five key zip codes in South Brooklyn, voiced as early as September 29 [11], may have contributed to a preexisting downward trend.

What’s more, a demonstrated narrow effect on a specific endpoint such as restaurant visits does not necessarily imply that the regulatory scheme had an overall deterrent effect on SARS-CoV-2 transmission in South Brooklyn. The COVID-19 incidence data in Fig. 4 do show a temporary decline during the weeks ending October 17, 24 and 31, after the regulatory scheme had entered into force. However, as in our interpretation of the restaurant visitation data in Fig. 7, COVID-19 incidence was declining during the week before any controls went into effect. And in view of the approximate 5-day incubation period between initial infection and the subsequent development of symptoms warranting testing [34], the incidence of the disease was likely to have been declining even earlier. These considerations reinforce the conclusion that the actual effect of the Governor’s regulatory scheme is, strictly speaking, not identifiable from the available data.

Even if the Governor’s regulatory scheme was at least partly responsible for retarding the surge of COVID-19 in the central red zone during the last three weeks of October, it is evident that the program’s success was short-lived. As Figs. 4 and 6 show, the decline in COVID-19 incidence during October was ultimately reversed by a wave of increasing disease incidence that had similarly overtaken the surrounding orange and yellow zones by the end of November. These observations beg the question: Why did the Governor’s scheme ultimately fail?

### Overreliance on Test Positivity

One clue is offered by the striking discordance between the upward surge of disease *incidence* and the continuing downward trend in *test positivity* during the week ending November 9, as shown in Fig. 5. The underlying explanation for the discordance of these two trends was the continuing expansion of testing, which diluted the rising incidence with an abundance of negative test results. It was just at this juncture that regulators, overly fixated on the test positivity rate, cut the size of the red zone in half and then converted the red zone to orange. Under this interpretation of the evidence, the Governor’s regulatory scheme did have an initial retardant effect on COVID-19 incidence, but subsequent premature withdrawal of regulatory controls neutralized the effect of the initial policy.

### Access Controls Versus Restrictions on Mobility

The regulations imposed in the Governor’s concentric regulatory zones were fundamentally controls on access – to eating places, to school buildings, to houses of worship, and to large meetings. They differ from the classic remedy of containment, which entails restrictions not just on access, but also on overall mobility [35-37]. Although restricting access to some critical locations is indeed likely to reduce disease propagation [38, 39], the question here is whether focused controls on access alone were sufficient to alter the underlying mobility patterns that served as the template for a surge in COVID-19 cases. While some have cited increasing indoor activity with the arrival of colder fall weather [40] or a trend toward large family gatherings as the Thanksgiving holiday approached [41], we have in mind more fundamental, well-established contact networks.

The hypothesis that the underlying mobility patterns within the regulated area in South Brooklyn were insufficiently altered by the Governor’s regulatory controls on access is supported by the stability of the interzone movement matrices before and after the promulgation of the regulatory scheme (Supplement Appendix Table A1) as well as absence of any significant change in the proportion of devices with no movements (Supplement Appendix Fig. A3).

### High-Mobility ZCTAs as Drivers of the Local COVID-19 Surge

One interpretation of the trends in Fig. 4 is that the surge in COVID-19 cases that ultimately overran South Brooklyn was a citywide phenomenon, and that the incidence of the disease was simply increasingly uniformly across all ZCTAs. However, the patterns of declining and rising COVID-19 incidence seen in Fig. 6 go against this interpretation. Once the incidence reached a low point around the week ending October 17, the subsequent increase was driven by ZCTAs along the southern and western borders of the area. During the resurgence of COVID-19 incidence, mobility patterns were hardly uniform, as shown in Fig. 8. In fact, the geospatial analysis demonstrates that five high-mobility ZCTAs – of which only two were part of the original red zone – were the main drivers of the resurgence in COVID-19, with a reproductive number close to 2, as shown in the panel labeled Inhomogeneous Mixing B1 in Fig. 9. That mobility was the critical determinant is further supported by the finding of the inferior performance of Model B2, based upon the Governor’s original partitioning of regulatory zones (Supplement Appendix Table C1).

### Conclusions

Our study is fundamentally observational. We did not analyze a macro-experiment in which various communities were randomly assigned to different regulatory controls or no intervention at all [42]. Still, our setup has many of the features of a natural experiment. The Governor’s announcement of a new regulatory regime on October 6, to become effective by October 9, could reasonably be characterized as an abrupt shock [7]. In view of the Mayor’s earlier threats to impose controls on certain zip codes in South Brooklyn [11], however, it could hardly be considered an unanticipated shock. The implementation of distinct regulations within each of three concentric regulatory zones provided natural intervention and control groups, and the results of our paired restaurant analysis (Fig. 7) suggest that the regulations were effective and enforced. While the restaurants stayed put, however, Supplement Appendix Table A1 and Fig. 8 show that the experimental participants crossed over from one zone to another. Midway through the intervention, on November 9, the Governor cut the regulatory zones in half [17], thereby contaminating the original experimental assignments.

Despite these flaws in design, we can nonetheless draw some reasonable inferences from the accumulated evidence. First, test positivity as a real-time indicator of regulatory effectiveness is fraught with potential biases [43, 44]. Here, the Governor and his advisors may have been led astray by a test positivity rate that was kept misleadingly close to 3 percent by an endogenous increase in testing among COVID-negative persons (Fig. 5).

Second, while restrictions on access to eating establishments and other high-risk venues may be narrowly effective (Fig. 7), they do not prevent people from moving around (Supplement Appendix Table A1, Fig. 8). In highly populous areas such as South Brooklyn, a halfway strategy of concentric regulatory zones based solely on access restrictions may be no substitute for the classic approach of concentric containment/quarantine areas [6, 37].

Third, overreliance on static rather than dynamic measures of disease burden to draw the boundaries of regulatory zones can prove to be highly misleading. Our geospatial analysis of COVID-19 incidence, entailing a dynamic model of COVID-19 incidence across 21 zip code tabulation areas (Fig. 9), identified five high-mobility ZCTAs where the reproductive number approached 2. Two of the five were not in the original red zone. Concentric zones may appear to be an effective regulatory approach in principle, but only if the boundaries are drawn correctly.

Fourth, policies restricting mobility can take many forms, including controls on transportation networks. There is substantial evidence pointing to the initial widespread dissemination of SARS-CoV-2 via New York City’s subway-based network during February-March 2020, followed by percolation of new infections within local community hotspots [45]. A policy of running express lines with limited density might have been an alternative to the complete shutdown of subway lines adopted in Wuhan [46].

Finally, in extreme cases, it may be necessary to impose stay-at-home restrictions. During an outbreak in September 2020 on the campus of the University of Wisconsin-Madison, the university administration barred students from leaving two highly infected residence halls. By the end of the month, COVID-19 incidence on campus had fallen below that of the surrounding county [47]. Even so, such stay-at-home orders may prove incompletely effective when disease propagation is dominated by intrahousehold transmission, as it was during the winter COVID-19 surge in Los Angeles County, a region with a high prevalence of multi-generational households [48]. Whether such a stay-at-home order would have been effective or even feasible in the case of South Brooklyn remains an open question.

## Data Availability

All programs and data are available at OSF Project "Brooklyn Concentric Zones 2020," https://osf.io/rquyx/

## Author Declarations

### Authorship

JEH is the sole author of this work. He is responsible for the conceptualization of the study, the data analysis, the drafting of the manuscript, and the creation of the figures.

## Acknowledgments

This article represents the sole opinion of its author and does not necessarily represent the opinions of the Massachusetts Institute of Technology, Eisner Health, or any other organization.

## Human Studies Approval

This study relies exclusively on publicly available data that contain no individual identifiers. No human studies approval was required.

## Public Posting of Data and Programs

Supporting programs and data have been posted at https://osf.io/rquyx/.

