## Supplement for "Failure of Concentric Regulatory Zones to Halt the Spread of COVID-19 in South Brooklyn, New York: October-November 2020"

### Appendix A

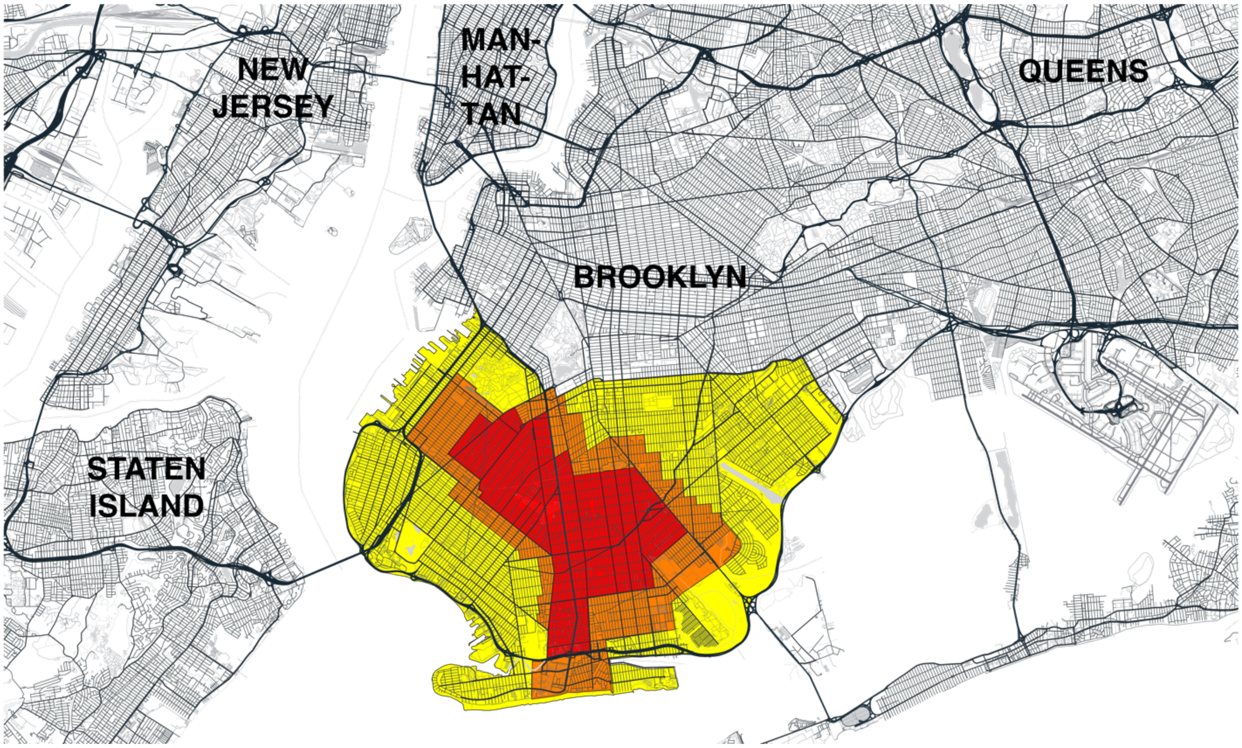

**Fig A1. Concentric Regulatory Zones Established in South Brooklyn, Overlaid on Street Map of the New York City Area.** The original three zones, color-coded red, orange, and yellow, went into effect on October 9, 2020 [1]. On October 21, the orange zone was redesignated as a yellow zone, while the red zone remained unchanged [2]. The entire regulated area, situated in South Brooklyn, was bounded along the north by New York State Route 27, originating in the northwest at exit 24 of Interstate I-278, with stretches along Prospect Parkway, Caton Avenue, Linden Boulevard, and then further bounded by Pennsylvania Avenue at the northeast end. Base map and data from New York City Planning [3].

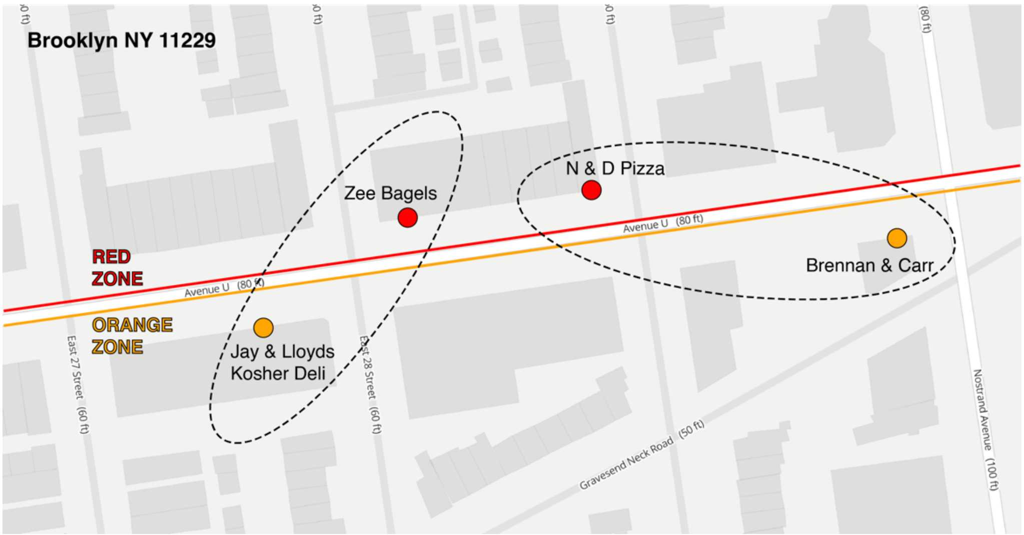

**Fig. A2. Illustrative Paired Points of Interest Overlapping the Boundary Between the Original Red and Orange Zones.** Avenue U bounds the red zone to the north and the orange zone to the south. *Zee Bagels* in the red zone was paired with *Jay & Lloyds Kosher Deli* (now permanently closed) in the orange zone. The distance between the two POIs was 61 meters. Similarly, *N & D Pizza* in the red zone was paired with *Brennan & Carr* in the orange zone. The distance between the two POIs was 92 meters. Base map and data from New York City Planning [4].

Table A1. Distribution of Device Movements Three Weeks Before and Three Weeks After the Establishment of Concentric Regulatory Zones on October 9, 2020 [a]

| Origin Zone | Percentage of Device Stops by Destination | | | | | Total Stops [d] |
|---|---|---|---|---|---|---|
| | Red | Orange | Yellow | Metro [b] | Outside [c] | |
| *Before (September 18 – October 8, 2020)* | | | | | | |
| Red | 57.2 | 9.8 | 9.4 | 16.8 | 6.8 | 479,899 |
| Orange | 7.6 | 56.7 | 13.1 | 17.7 | 4.9 | 577,847 |
| Yellow | 3.9 | 6.8 | 64.0 | 21.3 | 4.0 | 1,191,614 |
| *After (October 9 – October 29, 2020)* | | | | | | |
| Red | 57.6 | 9.4 | 9.2 | 16.8 | 7.0 | 487,557 |
| Orange | 7.1 | 56.8 | 12.7 | 18.4 | 5.0 | 576,975 |
| Yellow | 3.6 | 6.5 | 64.8 | 21.2 | 3.9 | 1,184,063 |

a. Each mobile device movement represented a stop in a particular census block group (CBG) for >1 minute. Each such stop had an origin and a destination CBG [5]. The table accounts only for movements of devices originating in the original red, orange, and yellow containment zones, where CBGs were classified according to the scheme in Fig. 3a. The percentages in each row add to 100%. The median dwell times (in minutes) for stops outside the home during September 18 – October 8 were: Red, 47; Orange, 35; and Yellow 42. The corresponding median dwell times during October 9 – 29 were: Red, 42; Orange, 29; and Yellow, 36.
b. Metro refers to destination CBGs in the remainder of the 17-county metropolitan region outside the red, orange, and yellow zones. This 17-county region included: one county in Connecticut (Fairfield); 7 counties in New Jersey (Bergen, Essex, Hudson, Middlesex, Monmouth, Passaic, Union); and 9 counties in the rest of New York City and New York state (Bronx, Kings, Nassau, New York, Queens, Richmond, Rockland, Suffolk, Westchester).
c. Outside refers to destination CBGs beyond the 17-county Metro region.
d. Total stops include all device stops originating from each zone during each 3-week period.

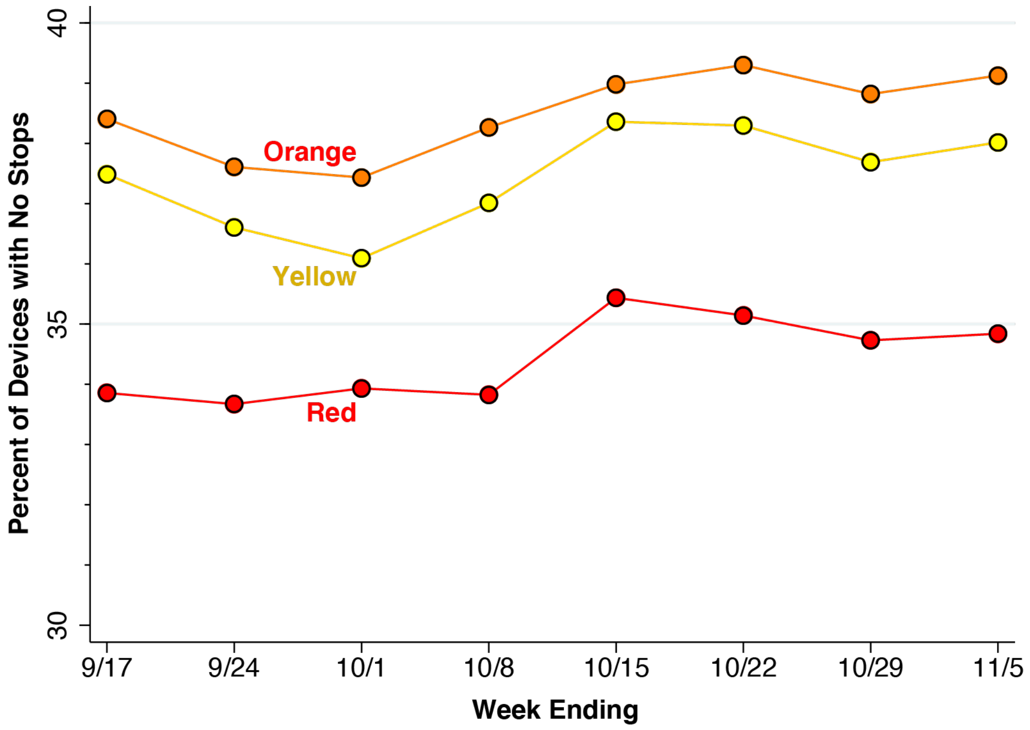

**Fig. A3. Proportion of Devices Homed in Each of the Original Zones That Made No Stops During Weeks Ending 9/17 – 11/5/2020.** Census block groups (CBGs) were mapped into regulatory zones in accordance with the scheme in Fig. 3a in the main manuscript. In each CBG during each day, we determined the number of devices making no stops, derived from the variable *bucketed_distance_traveled*, and the total number of devices, derived from the variable *device_count*, as recorded in the Safegraph social distancing database [5].

### Appendix B: Geospatial Model

*Homogeneous Mixing*

An area consists of a fixed number of geographically contiguous zones, indexed $k = 1,\ldots,K$. A total of $N_k > 0$ individuals reside in zone $k$. At the start of each time period $t = 1,\ldots,T$, each individual residing in each zone $k$ either remains within her zone of residence or temporarily moves to any one of the other $K - 1$ zones. At the end of time period $t$ and before the start of time period $t + 1$, the individual returns home to zone $k$. Let $n_{k\ell t} \geq 0$ denote the number of movements of individuals from zone $k$ into zone $\ell$ during time period $t$. Fig. B1 illustrates the types of movements within and between zones in a two-zone area at time $t$. All individuals' movements are accounted for, so that $\sum_\ell n_{k\ell t} = N_k$. Note that the number of individuals temporarily situated in zone $\ell$ during time period $t$ is $M_{\ell t} = \sum_k n_{k\ell t} \geq 0$.

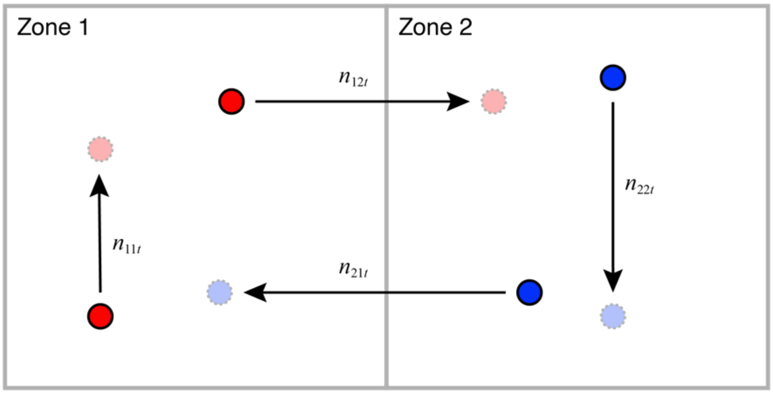

**Fig. B1. Four types of movements within and between zones in a two-zone area at time $t$.** Individuals represented by red circles reside in Zone 1 and individuals represented by blue circles reside in Zone 2. During time period $t$, $n_{11t}$ residents of Zone 1 stay within their home zone, while $n_{12t}$ residents of Zone 1 temporarily move to Zone 2. Similarly, $n_{22t}$ residents of Zone 2 stay within their home zone, while $n_{21t}$ residents of Zone 2 temporarily move to Zone 1. During period $t$, a total of $M_{2t} = n_{12t} + n_{22t}$ individuals are temporarily situated in Zone 2. Of these $M_{2t}$ individuals, the proportions originating from Zones 1 and 2, respectively, are $w_{12t} = n_{12t}/M_{2t}$ and $w_{22t} = n_{22t}/M_{2t}$. A total of $N_1 = n_{11t} + n_{12t}$ individuals permanently reside in Zone 1. Of these $N_1$ individuals, the proportions moving to Zones 1 and 2, respectively, are $v_{11t} = n_{11t}/N_1$ and $v_{12t} = n_{12t}/N_1$.

An infectious disease is spread though contact between individuals temporarily situated within the same zone. The disease has a latency, an incubation interval, and a duration of infectivity of exactly one time period. That is, an individual infected through contact with an infectious person during time period $t$ – 1 is diagnosed with the disease and herself becomes infectious to others only during time period $t$.

We keep track of the disease according to individuals' zone of residence $k$ and date of diagnosis $t$. We let $y_{kt} \geq 0$ denote the fraction of residents of zone $k$ who were diagnosed during time period $t$. We refer to these quantities as the incidence of the disease. The initial incidence $y_{k1}$ thus represents the fraction of individuals residing in zone $k$ who are diagnosed with the disease and are infectious to others during time period $t$ = 1. Except for these initially infectious individuals, the remainder of the population is susceptible to infection. We assume $y_{k1} \ll 1$, so that the initial proportion of susceptible individuals in each zone $k$ is approximately equal to 1.

We further assume homogeneous mixing of infectious and susceptible individuals. Let $w_{k\ell t}$= $n_{k\ell t}/M_{\ell t}$ represent the fraction of all individuals temporarily situated in zone $\ell$ during time period $t$ who reside in zone $k$. Then the conditional probability of infection of a randomly selected, susceptible person temporarily situated in zone $\ell$ during time period $t$ is $p_{\ell t}$ = $\alpha \sum_r w_{r\ell t} y_{rt}$, where $\alpha$ > 0 is the reproductive number, an indicator of the efficiency of transmission. (In this expression, we have used the summation index $r$ for zones of residence, rather than $k$.)

Now consider a randomly selected, susceptible individual residing in zone $k$. The probability that this individual will be temporarily situated in zone $\ell$ during time period $t$ is $v_{k\ell t}$ = $n_{k\ell t}/N_k$. Hence, the unconditional probability that this resident of zone $k$ will be infected via contact with an infectious person during time period $t$ is $\sum_\ell v_{k\ell t}\, p_{\ell t}$ or equivalently, $\sum_\ell (v_{k\ell t})\, (\alpha \sum_r w_{r\ell t} y_{rt})$. We thus have the dynamic equation of motion of the incidence of infection in each zone in each time period:

$$y_{k,t+1} = \alpha \sum_\ell \sum_r v_{k\ell t} w_{r\ell t} y_{rt} \tag{B1}$$

Equation (B1) is a linear system of difference equations, in which the incidence of new cases $y_{k,t+1}$ diagnosed during time period $t$ + 1 among residents of zone $k$ depends on the incidence $\{y_{rt}\}$ of cases previously diagnosed at time $t$ in all zones $r$ = 1,…,$K$.

It will be useful to convert equation (B1) into matrix notation. For each time period $t$, let $V_t$ be the $K \times K$ square matrix with typical element $v_{k\ell t}$= $n_{k\ell t}/N_t$, which, as noted above, represents the fraction of all residents of zone $k$ who temporarily move to zone $\ell$ during time period $t$. The elements in the rows of $V_t$ sum to 1. Let $W_t$ be the $K \times K$ square matrix with typical element $w_{k\ell t}$= $n_{k\ell t}/M_{\ell t}$, which, as noted above, represents the fraction of all individuals temporarily situated in zone $\ell$ during time period $t$ who reside in zone $k$. The elements of the columns of $W_t$ sum to 1. Finally, let $Y_t$ denote the $K \times 1$ column vector $(y_{1t}, \dots, y_{Kt})'$. Then our dynamic model can be represented in matrix form as:

$$Y_{t+1} = \alpha\, V_t\, W_t{}'\, Y_t \tag{B2}$$

In equation (B2), all the elements of the $K \times 1$ column vector $V_t\, W_t{}'\, Y_t$ are multiplied by the same scalar parameter $\alpha$. If we already have data on the movements $\{n_{k\ell t}\}$ between zones and data on the incidence $\{y_{kt}\}$ of the disease, then the matrices $V_t$ and $W_t$ and the vectors $Y_t$ are likewise observed quantities, and thus we can estimate the reproductive number $\alpha$ as the only unknown parameter.

*Inhomogeneous Mixing*

We consider two modifications of equation (B2) to capture inhomogeneous mixing [6]. First, movements of individuals who remain within their zone of residence may have a lower reproductive number $\alpha_0$. Second, movements of individuals who reside in or enter certain high-risk zones may have a higher reproductive number $\alpha_1$. All other movements are assumed to have the same reproductive number $\alpha_2$.

To capture within-zone movements, let $D_t$ = diag($V_t W_t{}'$) denote the $K \times K$ square matrix with the same diagonal elements as $V_t W_t{}'$ but zero off-diagonal elements. Define the $K \times K$ square matrix $X_t = V_t W_t{}' - D_t$, that is, the matrix $V_t W_t{}'$ with its diagonal elements set to 0. Then our model becomes:

$$Y_{t+1} = \alpha_0 D_t Y_t + \alpha_1 X_t Y_t \qquad \text{(B3)}$$

To further capture differences in transmission efficiency between zones, we partition the set of zones into two nonempty subsets, $L$ and $H$, representing the low- and high-transmission zones, respectively. The former has $K_L$ elements and the latter has $K_H = K - K_L$ elements. We assume all zones in $L$ have the same reproductive number $\alpha_1$, while all zones in $H$ have reproductive number $\alpha_2$. Partition the square $K \times K$ matrix $X_t$ vertically into two matrices: a $K \times K_L$ matrix $X_{tL}$, and a $K \times K_H$ matrix $X_{tH}$. Similarly, partition the column vector $Y_t$ horizontally into two vectors: $Y_{tL}$ and $Y_{tH}$. Then equation (B3) becomes:

$$Y_{t+1} = \alpha_0 D_t Y_t + \alpha_1 X_{tL} Y_{tL} + \alpha_2 X_{tH} Y_{tH} \qquad \text{(B4)}$$

*Model Implementation*

The area under study consists of $K$ = 21 ZCTAs situated in South Brooklyn, as shown in Fig. 2 of the main text. We focus on the 7-week period running from the week ending October 17 through the week ending November 28, that is, the last 7 images in Fig. 6 in the main text. We derive the incidence $y_{kt}$ for $k$ = 1,...,21 and $t$ = 1,...,7 from confirmed COVID-19 cases per 100,000 population, as reported by the New York City Department of Health [7].

We relied upon the variables variables *origin_census_block_group* and *destination_cbgs* in the Safegraph Social Distancing database [5] to identify all device movements between census block groups (CBGs) within the 21-ZCTA area. Assigning each CBG to a specific ZCTA based upon the location of its centroid, we were thus able to compile data $n_{k\ell t}$, representing the number of device movements from ZCTA $k$ to ZCTA $\ell$ during week $t$. We further relied on the variable *bucketed_distance_traveled* to count the number of devices that made no movements in each ZCTA $k$ during each week $t$, and then added these counts to the within-ZCTA movements $n_{kkt}$. While we also observed device movements beyond the 21-ZCTA regulated area, as well as movements into the regulated area from outside, we focused sharply on the regulated area in order to ascertain how the traffic between local ZCTAs influenced the dynamics of COVID-19 transmission.

Given the incidence data $\{y_{kt}\}$ and movement counts $\{n_{k\ell t}\}$, we estimated the models of equations (B2), (B3) and (B4) by weighted least squares, where the weights were the populations of each ZCTA.

### Appendix C

**Table C1. Parameter Estimates for Geospatial Models with Homogeneous and Inhomogeneous Mixing.** [a]

| *Parameter* [b] | *Homogeneous Mixing, Eq. (2)* [c] | *Inhomogeneous Mixing, Eq. (3)* [c] | | |
|---|---|---|---|---|
| | | *Model A* [d] | *Model B1* [d] | *Model B2* [d] |
| $\alpha$ | **1.157** (0.026) | | | |
| $\alpha_0$ | | 1.112 (0.118) | 1.099 (0.115) | 1.120 (0.117) |
| $\alpha_1$ | | 1.228 (0.187) | 0.846 (0.236) | 0.735 (0.369) |
| $\alpha_2$ | | | **1.910** (0.324) | 1.485 (0.249) |
| $R^2$ statistic | 0.942 | 0.942 | 0.945 | 0.943 |
| Root MSE | 23.846 | 23.928 | 23.416 | 23.794 |
| p-value [d] $H_0: \alpha_0 = \alpha_1$ | | 0.699 | | |
| p-value [d] $H_0: \alpha_1 = \alpha_2$ | | | 0.012 | 0.124 |

a. All models had 126 observations. Root MSE = Root mean squared error.
b. Parameters described in equations (2), (3) and (4) in main text and in equations (B2) – (B4) above.
c. Standard error is shown below each parameter estimate. Estimate is shown in bold-face if two-sided null hypothesis that parameter = 1 is rejected (based upon F-test) at $p < 0.05$.
d. Model A distinguished between within-ZCTA effects ($\alpha_0$) and all other effects ($\alpha_1$). Models B1 and B2 distinguished between within-ZCTA effects ($\alpha_0$), other low-risk effects ($\alpha_1$), and other high-risk effects ($\alpha_2$). Model B1 was based on the classification of five ZCTAs (11223, 11229, 11235, 11224, and 11214) as high risk, while Model B2 was based upon classification of all original red-zone ZCTAs as high risk.
e. Other tests of two-sided null hypotheses ($H_0$) based upon F-tests.
